# Early economic modelling of a new pharmacotherapeutic treatment pathway for children with monogenic obesity

**DOI:** 10.64898/2026.05.13.26353098

**Authors:** Padraig Dixon, Helen Stewart, Olu Onyimadu, David B.N. Lim, Nikki Davis

## Abstract

**Background:** Early onset obesity in children, almost always accompanied by significant health complications, may be driven by rare genetic variants that influence appetite, metabolism, and nutrient absorption. Traditional treatment approaches are usually insufficient for those with monogenic obesity of this type. Glucagon-like peptide-1 (GLP-1) receptor agonists, such as semaglutide, and related drugs such as melanocortin 4 receptor agonists, have emerged as promising first-line treatments for severe obesity. There is no established protocol or pathway in England for identifying children with monogenic obesity who could benefit from these and similar treatments

**Methods:** We undertook early economic modelling to examine the cost-effectiveness, from a health service perspective, of implementing a new pharmacotherapeutic care pathway for the identification and treatment of monogenic obesity in children. We modelled a hypothetical population of children with hyperphagia and body mass index (BMI) three standard deviations above mean values for age and sex. We evaluated the clinical decision to initiate the pathway using a decision tree model with patient quality-adjusted life years (QALYs) and NHS healthcare costs 12 months from an initial clinic visit as outcomes, and calculated incremental cost effectiveness ratios and a cost-effectiveness acceptability curve.

**Results:** Both costs and QALYs were higher under further investigation (£3,247 and 0.47 QALYs) compared to no further investigation (£1,589 and 0.24 QALYs). The incremental cost-effectiveness ratio in the base case was £7,133 per QALY. Further examination of these children was therefore likely to be cost effective in this model.

**Conclusion:** A decision-tree model suggested that further investigation of severely obese children potentially eligible for treatment with semaglutide is likely to be cost-effective for the NHS. However, this result is associated with uncertainty arising from a lack of evidence for many key model parameters.

## 1 Introduction

Variants in genes associated with the production of hormones that regulate appetite, metabolism, and nutrient absorption can disrupt the processes that control body weight (1–4). Children carrying these variants may experience severe early onset obesity, type 2 diabetes, high blood pressure, and heart disease (2, 4–6). Severely obese children typically have difficulty with physical activities and are also at a higher risk of developing psychological problems such as low self-esteem and depression (4, 6–8).

Recently developed agonists of the glucagon-like peptide-1 (GLP-1) receptor, which plays a central role in the regulation of appetite and satiety, offer considerable promise as a first-line treatment for obesity (9–12). Dual incretin receptor agonists, such as tirzepatide, target both the GLP-1 receptor and GIP (glucose-dependent insulinotropic polypeptide) receptor and may offer further advantages beyond monotherapy for some patients.

Melanocortin-4 receptor (MC4R) agonists, such as setmelanotide, are another therapy class that may have particular relevance for children with very rare forms of monogenic obesity.

Genetic testing in severely obese children suspected to have early onset monogenic obesity could support more accurate diagnoses, guide treatment decisions, improve long-term health and psychological outcomes and enable children and their carers to reach more informed decisions about their care and their health (2, 4, 13, 14). Moreover, wider genetic testing following an initial diagnosis in the child proband, may also reveal family members who share or have an increased genetic risk for the same condition, potentially allowing them to benefit from treatment.

At present, there is no established care pathway in England to identify children who have or are suspected to have monogenic obesity that may be amenable to treatment. In this paper, we consider the initial economic case for a new pathway to identify children suspected of monogenic obesity and who may benefit from treatment with semaglutide, a GLP-1 agonist. Semaglutide enhances fasting and post-prandial insulin secretion, suppresses appetite, and slows gastric emptying (12, 15). Despite the lack of substantial evidence concerning the use of semaglutide in severely obese children, the need to identify treatment and management options for this group is critical. In other rare cases, children undergoing treatment may benefit from MC4R receptor agonists such as setmelanotide (16).

## 2 Methods

### 2.1 Introduction

We undertook early economic modelling to examine the cost-effectiveness, from a health sector/NHS perspective, of a testing pathway for children. We describe this modelling approach as “early” because evidence is still limited on a number of important parameters relevant to the decision to refer children to the testing pathway. The modelling approach combined available evidence with (where necessary) assumptions regarding model parameters to explore likely value for money and cost effectiveness ahead of more extensive prospective and interventional studies in this patient population.

We assumed these children would present to clinic with a body mass index (BMI) standard deviation score (SDS) greater than three for age and sex, accompanied by hyperphagia (as assessed by a tool such as the Dykens’ Hyperphagia Questionnaire (17)) and possibly presenting with severe early onset obesity. We modelled the decision to undertake further testing of children to assess their eligibility for treatment. Children for whom no further testing is undertaken at this initial decision point were assumed to receive standard care for severe obesity, which would involve paediatric endocrinology specialists and may also include dietetics, psychology, and family support.

We did not explicitly model the process by which children are referred to clinic. In practice, this could encompass a number of different paediatric and adult clinical specialities and services. Referrals could potentially also be made from “Complications from Excess Weight (CEW)” clinics (18), which bring together doctors, psychologists and others to support children and young people with complications arising from excess weight, or from the National Childhood Measurement Programme (19).

The proposed pathway is shown in Figure 1.

**Figure 1.**
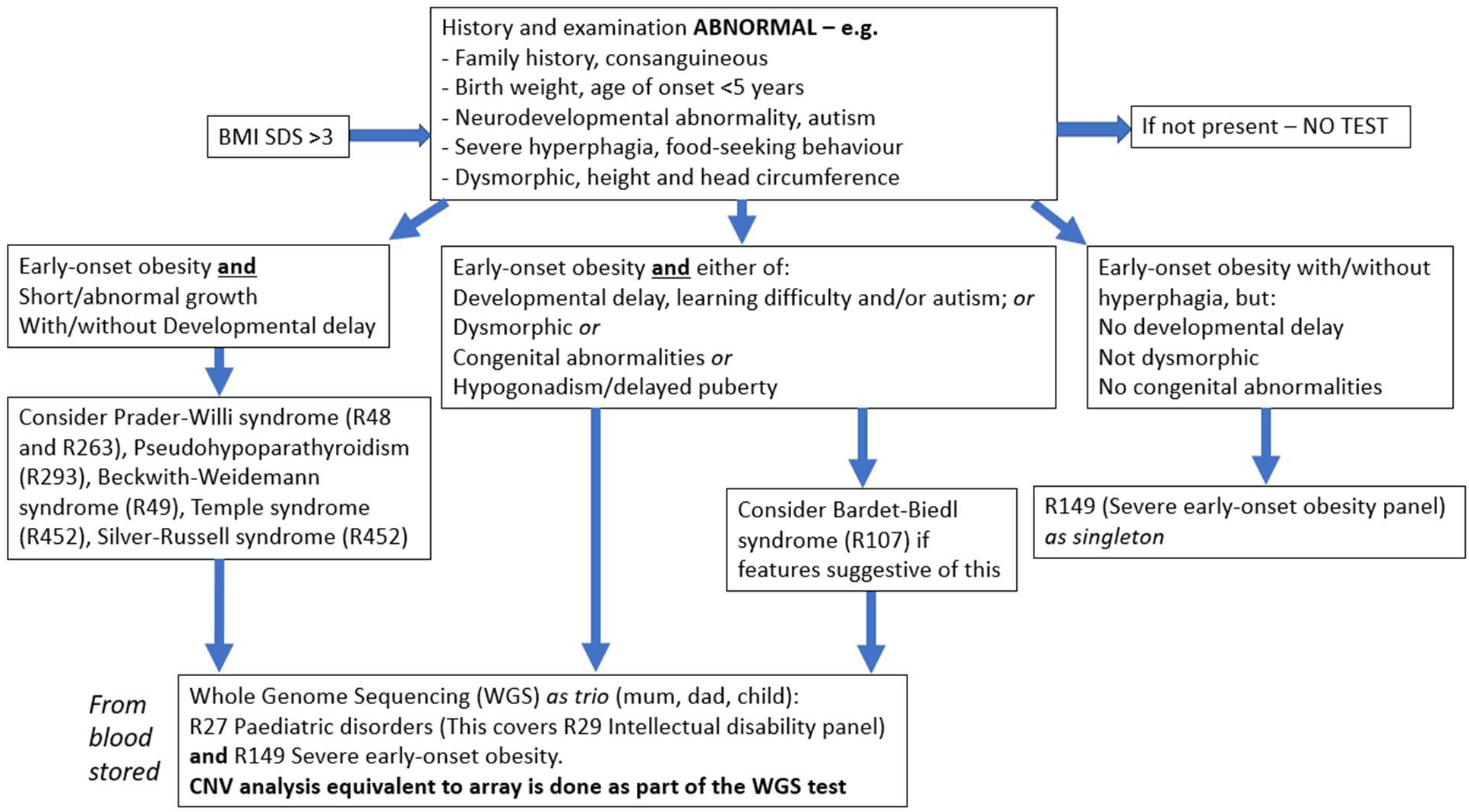
Clinical pathway

### 2.2 Decision analytic modelling

We modelled the choice between “no further examination” and further examination of these children using a decision tree model (20). Decision trees are an example of decision-analytic modelling, which enables the structured representation and modelling of outcomes and uncertainties linked to various alternative courses of action (21). This type of modelling is central to economic evaluation and health technology assessment, the primary objective of which is to compare the relative benefits of different treatment options, service provisions, or other interventions (22). Decision trees derive their name from their graphical representation, which resembles a branching structure. Each branch signifies distinct and mutually exclusive future possibilities.

In the context of clinical pathways, decision trees illustrate the sequential progression of discrete decisions, and their associated costs, and outcomes that occur in reasonably close temporal proximity along decision pathways. We used a one-year time horizon in the model. We recognize that the impacts of severe obesity and its resolution may have lifelong consequences. However, our focus in this analysis was on modelling the likely near-term impacts of initiating this pathway for severely obese children given the prospect of offering a substantial proportion of these children an effective therapy and given the lack of evidence on longer term outcomes in this patient group.

### 2.3 Decision tree model

We developed the decision tree to reflect the proposed clinical pathway for these children. The decision tree is represented in Figure 2.

**Figure 2.**
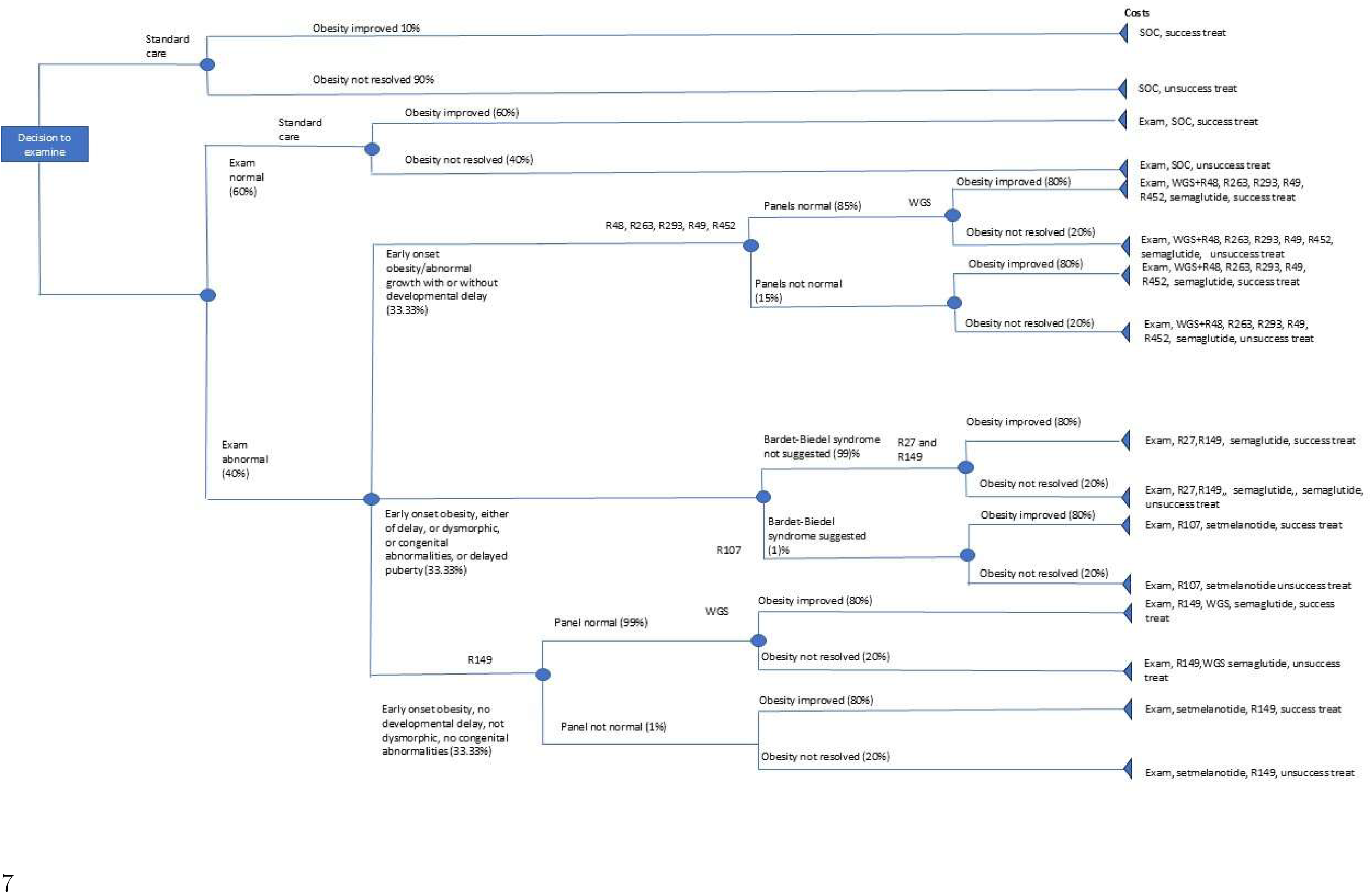
Decision tree of proposed pathway

The decision tree, which is read from left to right, starts with the initial clinical decision to further examine the child. If not examined, the child will continue usual care for their obesity, and will have a probability of improvement in the next 12 months. If examined, a number of further investigations may take place.

The square nodes represent decision points. The choice of which subsequent branch to take is made by the clinician in consultation with the patient. Only one of the alternative options (represented by different branches and subsequent treatment and outcome trajectories) may be chosen. The circular nodes represent random events and are always associated with a particular probability to quantify the uncertainty around these events. The triangular terminal nodes represent the final stage of a particular branch or path through the tree and at which the final outcomes are assessed. No further events or progress through the tree is permitted after a terminal node has been reached.

Outcomes at each terminal node are represented by values depicting both healthcare costs and quality-adjusted life-years (QALYs). Expressing outcomes in these terms enables the undertaking of a cost-effectiveness analysis, the details of which are described below. The costs and QALYs at each terminal node are the (cumulated) costs and QALYs from all nodes along a specific pathway through the decision tree. The outcomes of different pathways will therefore differ according to the decisions and chance events encountered before and upon reaching the pathway’s terminal node.

### 2.4 Cost-effectiveness modelling

We studied the cost-effectiveness of further examination compared to no further examination with standard care for children with a with hyperphagia and who may have early onset obesity. We used an NHS health sector perspective and a one-year time horizon for the analysis in which neither costs nor effects were discounted. Costs are reported in 2023-2024 UK sterling price levels. Quality-adjusted life years (QALYs) were used as the measure of effects, for which we assumed no deaths during the period of 12-month follow-up after the initial examination. We relied on published utility values to characterise quality-of-life outcomes in these children without assuming or relying on the use of a particular quality-of-life instrument.

The most cost-effective decision (further examination or no further examination) is, in principle, the most appropriate intervention to implement, given current information about the parameters (such as the effectiveness and tolerability of semaglutide in this population) and conditional on model structure (such as the number and types of decisions modelled). However, there is inherent uncertainty regarding the parameters with which this model is populated, including the probabilities associated with each chance node, as well as the cost and QALY outcomes associated with the terminal nodes. We linked parameter values to probability distributions to represent the uncertainty associated with each parameter.

We then implemented probabilistic sensitivity analysis (21) in which the decision tree was simulated multiple (N=10,000) times. Each Monte Carlo simulation randomly sampled values from distributions associated with each model parameter. The core output of the probabilistic sensitivity analysis comprised the expected values (averaged over all N estimates from the simulation) of mean costs and mean QALYs with associated estimates of their uncertainty. We modelled all probabilities using beta distributions to obtain values in the interval [0,1]. Costs were either drawn from log normal distributions (costs associated with improved and unimproved obesity taken from Onyimadu (23)) or from normal and truncated (at £0 or some costs) normal distributions.

We used these outputs to calculate the net monetary benefit (22) of the initial decision to undertake further examination. Net monetary benefit (NMB) is defined as: NMB=ΔQALYs* `λ` − ΔCosts, where Δ is the incremental difference operator. In each case, this operator is applied to the difference between QALYs and costs associated with the initial decision regarding further examination. The lambda (`λ`) term represents the cost-effectiveness threshold, which reflects the rate at which the health system converts monetary inputs into health outputs. We focused on threshold values of £25,000 to £35,000 per QALY. We also calculated the cost-effectiveness acceptability curve, which indicates the probability that the decision to undergo further testing is cost effective for different values of the cost-effectiveness threshold. We also represented uncertainty around the point estimate of cost-effectiveness obtained from the probabilistic sensitivity analysis using a scatter plot on the cost-effectiveness plane.

The cost-effectiveness decision tree model was coded in R (version 4.3.3). The code and all data used in the model are available at github.com/pdixon-econ/mono-obesity-decision-tree.

## 3 Data

The model parameters and their associated distributions used to populate the decision tree represented in Figure 1 are described in Table 1. As our period of follow-up was assumed to be 12 months, we describe “improvements” in obesity rather than resolution, as in most if not all cases it is not realistic to expect a complete transition to normal weight and elimination of weight-related comorbidities after this length of time.

**Table 1.**
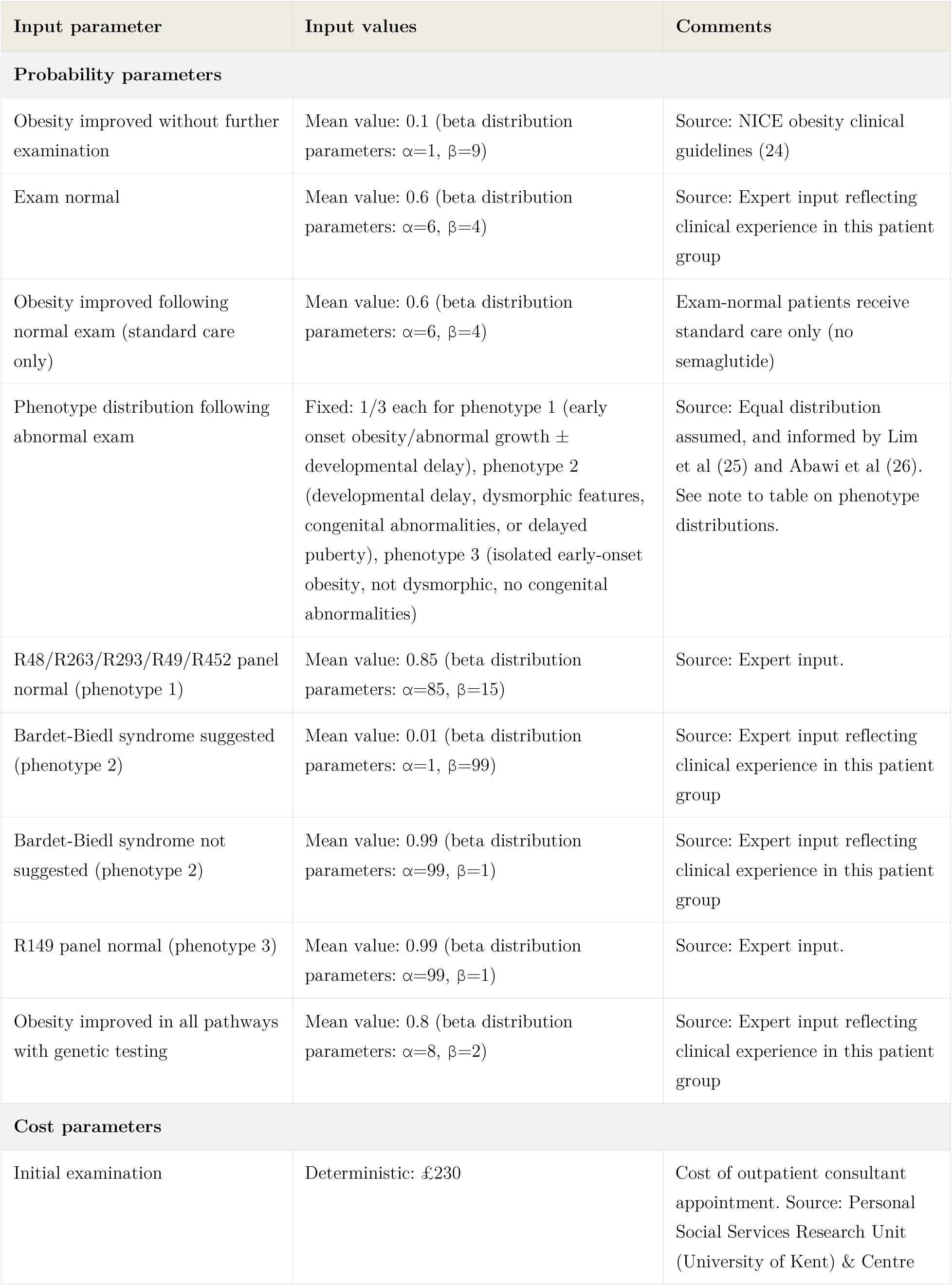
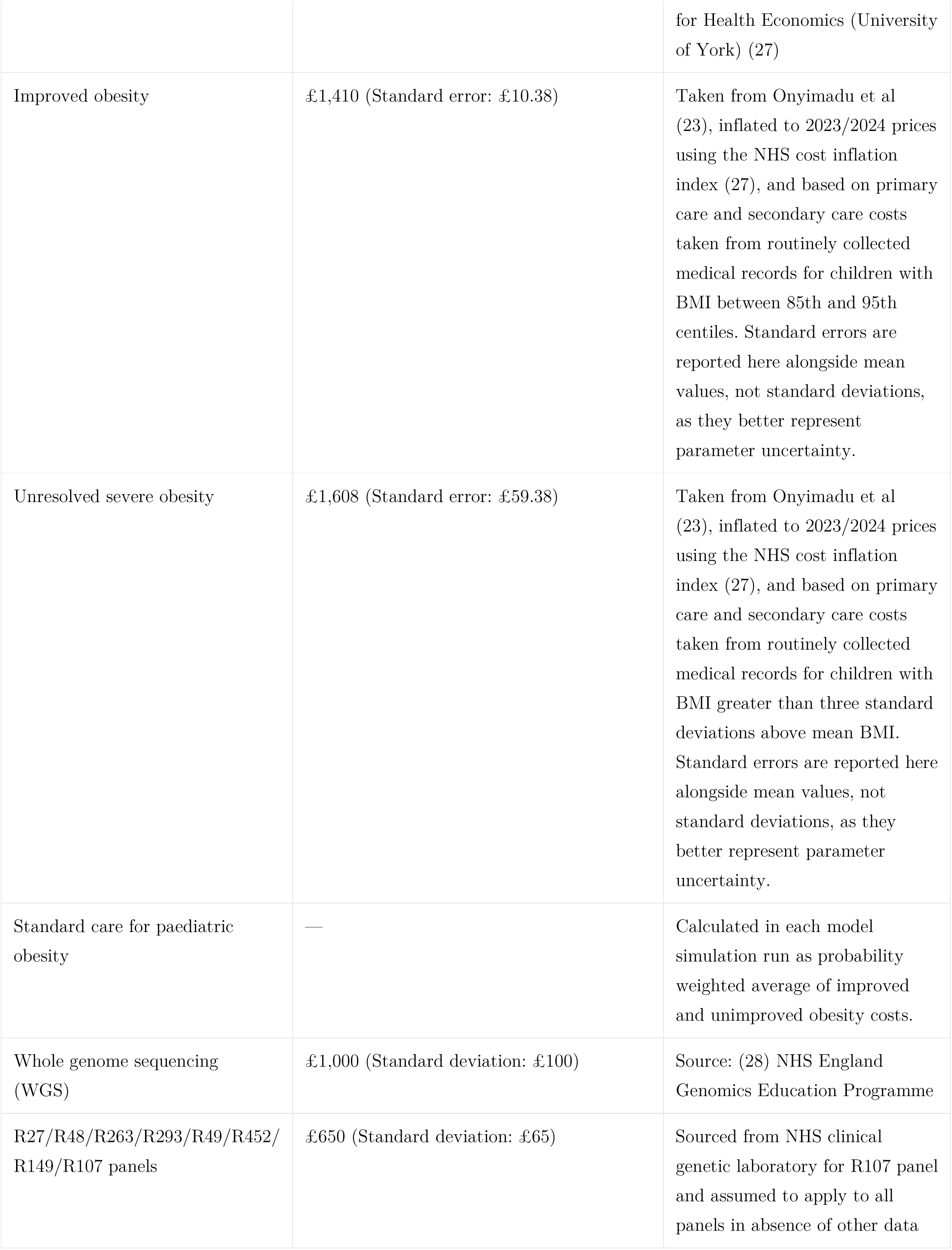
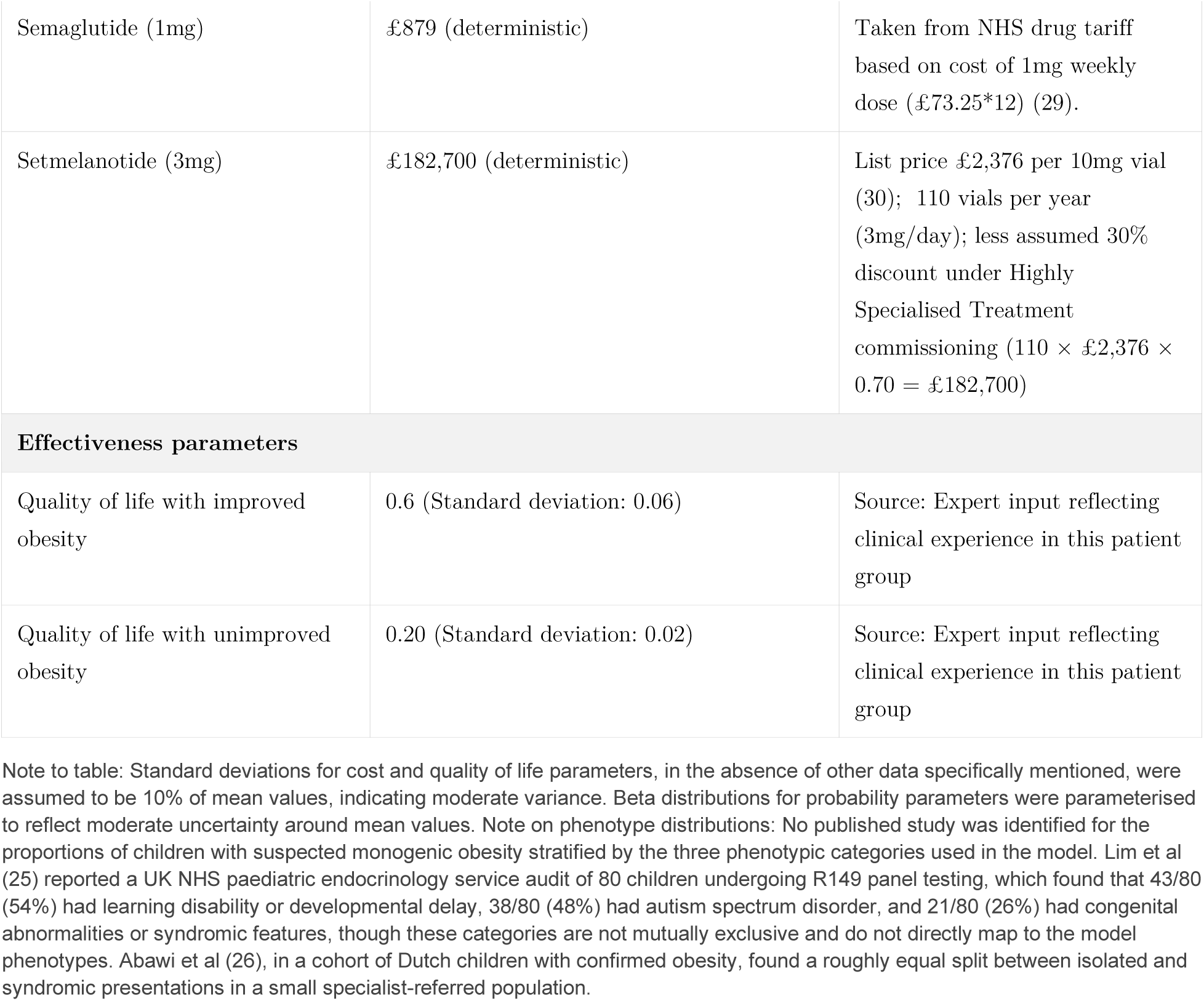
Input parameters used in decision tree.

| Input parameter | Input values | Comments |
| --- | --- | --- |
| **Probability parameters** |  |  |
| Obesity improved without further examination | Mean value: 0.1 (beta distribution parameters: $\alpha=1$, $\beta=9$) | Source: NICE obesity clinical guidelines (24) |
| Exam normal | Mean value: 0.6 (beta distribution parameters: $\alpha=6$, $\beta=4$) | Source: Expert input reflecting clinical experience in this patient group |
| Obesity improved following normal exam (standard care only) | Mean value: 0.6 (beta distribution parameters: $\alpha=6$, $\beta=4$) | Exam-normal patients receive standard care only (no semaglutide) |
| Phenotype distribution following abnormal exam | Fixed: 1/3 each for phenotype 1 (early onset obesity/abnormal growth ± developmental delay), phenotype 2 (developmental delay, dysmorphic features, congenital abnormalities, or delayed puberty), phenotype 3 (isolated early-onset obesity, not dysmorphic, no congenital abnormalities) | Source: Equal distribution assumed, and informed by Lim et al (25) and Abawi et al (26). See note to table on phenotype distributions. |
| R48/R263/R293/R49/R452 panel normal (phenotype 1) | Mean value: 0.85 (beta distribution parameters: $\alpha=85$, $\beta=15$) | Source: Expert input. |
| Bardet-Biedl syndrome suggested (phenotype 2) | Mean value: 0.01 (beta distribution parameters: $\alpha=1$, $\beta=99$) | Source: Expert input reflecting clinical experience in this patient group |
| Bardet-Biedl syndrome not suggested (phenotype 2) | Mean value: 0.99 (beta distribution parameters: $\alpha=99$, $\beta=1$) | Source: Expert input reflecting clinical experience in this patient group |
| R149 panel normal (phenotype 3) | Mean value: 0.99 (beta distribution parameters: $\alpha=99$, $\beta=1$) | Source: Expert input. |
| Obesity improved in all pathways with genetic testing | Mean value: 0.8 (beta distribution parameters: $\alpha=8$, $\beta=2$) | Source: Expert input reflecting clinical experience in this patient group |
| **Cost parameters** |  |  |
| Initial examination | Deterministic: £230 | Cost of outpatient consultant appointment. Source: Personal Social Services Research Unit (University of Kent) & Centre |
|  |  | for Health Economics (University of York) (27) |
| Improved obesity | £1,410 (Standard error: £10.38) | Taken from Onyimadu et al (23), inflated to 2023/2024 prices using the NHS cost inflation index (27), and based on primary care and secondary care costs taken from routinely collected medical records for children with BMI between 85th and 95th centiles. Standard errors are reported here alongside mean values, not standard deviations, as they better represent parameter uncertainty. |
| Unresolved severe obesity | £1,608 (Standard error: £59.38) | Taken from Onyimadu et al (23), inflated to 2023/2024 prices using the NHS cost inflation index (27), and based on primary care and secondary care costs taken from routinely collected medical records for children with BMI greater than three standard deviations above mean BMI. Standard errors are reported here alongside mean values, not standard deviations, as they better represent parameter uncertainty. |
| Standard care for paediatric obesity | — | Calculated in each model simulation run as probability weighted average of improved and unimproved obesity costs. |
| Whole genome sequencing (WGS) | £1,000 (Standard deviation: £100) | Source: (28) NHS England Genomics Education Programme |
| R27/R48/R263/R293/R49/R452/R149/R107 panels | £650 (Standard deviation: £65) | Sourced from NHS clinical genetic laboratory for R107 panel and assumed to apply to all panels in absence of other data |
| Semaglutide (1mg) | £879 (deterministic) | Taken from NHS drug tariff based on cost of 1mg weekly dose (£73.25*12) (29). |
| Setmelanotide (3mg) | £182,700 (deterministic) | List price £2,376 per 10mg vial (30); 110 vials per year (3mg/day); less assumed 30% discount under Highly Specialised Treatment commissioning ($110 \times £2,376 \times 0.70 = £182,700$) |
| **Effectiveness parameters** |  |  |
| Quality of life with improved obesity | 0.6 (Standard deviation: 0.06) | Source: Expert input reflecting clinical experience in this patient group |
| Quality of life with unimproved obesity | 0.20 (Standard deviation: 0.02) | Source: Expert input reflecting clinical experience in this patient group |

Note to table: Standard deviations for cost and quality of life parameters, in the absence of other data specifically mentioned, were assumed to be 10% of mean values, indicating moderate variance. Beta distributions for probability parameters were parameterised to reflect moderate uncertainty around mean values. Note on phenotype distributions: No published study was identified for the proportions of children with suspected monogenic obesity stratified by the three phenotypic categories used in the model. Lim et al (25) reported a UK NHS paediatric endocrinology service audit of 80 children undergoing R149 panel testing, which found that 43/80 (54%) had learning disability or developmental delay, 38/80 (48%) had autism spectrum disorder, and 21/80 (26%) had congenital abnormalities or syndromic features, though these categories are not mutually exclusive and do not directly map to the model phenotypes. Abawi et al (26), in a cohort of Dutch children with confirmed obesity, found a roughly equal split between isolated and syndromic presentations in a small specialist-referred population.

There was little to no published data available for most model parameters. Parameter estimates were in many cases based on the expert judgement and experiences of clinical and laboratory colleagues in providing care to these children.

### Probability parameters

There is little evidence on improvements in children with severe obesity with or without the use of GLP-1 agonists. NICE reported (31) in 2013 (before the GLP-1 era) that approximately 79% of children who are obese in early teens will remain obese as adults, 13 suggesting an upper bound of improvement of approximately 20% amongst children older than those modelled in this study and without (in most cases) the severe obesity and hyperphagia of the present study. In practice, clinical experience amongst the authorship group in treating this group suggests that, at most, some 10% might experience improved management in the 12 months after their first encounter with a weight management clinic, and we used this value in the model.

### Cost parameters

We assumed the initial consultation – at which the further examination decision is taken - was based on the NHS cost of an outpatient consultant appointment and was modelled as a deterministic cost. Based on clinical experience of the authorship group, we assumed that semaglutide would initially be dosed as 0.25mg per week, and gradually titrated to a higher dose (32). We assumed that the average dose over the 12 months of model follow-up would be 1mg based on clinical experiences. We obtained costs for semaglutide from the NHS drug tariff (29), and the costs of setmelanotide from (30). We assumed a mean cost of £1,000 for whole genome sequencing (28). We used a cost of £650 for the R149 panel test based on laboratory input. We assumed that this cost applied to other panels.

We used data from (23) to estimate costs associated with paediatric BMI. These data were sourced from primary care records in the Clinical Practice Research Datalink (CPRD) resource linked to Hospital Episode Statistics for Secondary care. CPRD contains routinely collected data from practices using EMIS Web® electronic patient record software and is representative of the wider NHS England population. Costs reflected all consultations and prescriptions in primary care, as well as any secondary care received via inpatient, outpatient or accident and emergency appointments in NHS settings.

Costs were measured as the mean annual costs for all children aged 5 to 10 years in the year after an index BMI measurement was made in primary care between 1 January 2015 and 30 March 2019. We measured these costs in children with BMI at a SDS score greater than 3 (for age and sex), as well as children in an overweight category. We assumed that children with “improved” obesity at the end of one year transitioned to overweight rather than healthy weight status. We defined overweight as the 85^th^ to 95^th^ percentiles of BMI. Costs were estimated using a two-part generalized linear model with gamma distribution and log-link function. These marginal effects represent the difference in costs between a specific category of BMI and the reference category of health weight, conditional on sex, age, ethnicity, region, and an index of multiple deprivation (IMD). These cost are similar between these categories, and were primarily driven by prescription costs for routine items. This is notwithstanding likely future divergences in healthcare costs between these groups of children as (further) comorbidities develop and treatment complexity increases.

### Quality of life parameters

There is a paucity of evidence on quality-of-life measurement in children with severe obesity. The mean utility for children of healthy weight reported in the Brown et al (33) meta-analysis of 0.85. This figure was obtained from a meta-analysis of studies reporting primary data collection of utility values from children, with results reported by weight status in paediatric populations (mean or median target age ≤ 18 years), written in the English language, and published up to May 2017. The Brown et al study found minimal impact of overweight and obesity on utility scores. These results do not appear to capture the full impact of severe obesity experienced by the type of children included in our analysis. In particular, these children tend to present to clinic with complex patterns of comorbidity.

Kwon et al (34) found substantial decrements to utility associated with these and other typical comorbidities in children. The modelling work Wiedemann et al (35) indicated severe adverse mortality and comorbidity profiles in children with severe early onset obesity, highlighting the significant quality of life burden likely to be experienced by these children. This analysis and clinical experience both suggest that the minimal impact of obesity reported in Brown et al (33) is unlikely to reflect the quality-of-life impacts associated with the severely obese children that are the subject of the present study.

However, the exact extent of these comorbidities, and potential additive or multiplicative effects of multiple comorbid conditions on utility, are not reported for this patient group.

We therefore used an arbitrary but possibly conservative utility value of 0.20 for children in this category. This is approximately one-quarter of the quality-of-life reported by Brown et al for children with normal weight (0.85). We also used an arbitrary value of 0.60 for children’s quality of life at 12 months following successful treatment of severe obesity. This recognises that these children will experience an improvement in weight, and likely also in the burden of comorbid conditions, but nevertheless are still likely to be obese and experiencing a quality-of-life below their non-obese peers.

For instance, van Boxel et al’s (32) small cohort study focused on children aged 10 to 18 years with a BMI above 35 kg/m² and at least one weight-related comorbidity, who were treated with semaglutide. Among the 14 children with 12 months of data available, the study found an average BMI reduction of 10 kg from a mean baseline weight of 108kg.

This amounted to a reduction in the prevalence of excess weight (calculated in relation to target weight as a function of height, age and gender) of approximately 18%.

We also undertook one-way deterministic sensitivity analysis for this parameter as follows. We first assumed that children with “improved” obesity would attain mean quality of life reported for children of normal weight in Brown et al (33) of 0.85, rather than the 0.60 reported in the base case analysis. This gives a high upper bound on what an effective obesity therapy could theoretically achieve in this group of children. In the second deterministic sensitivity analysis, we suppressed the difference between QALYs for the “improved” and “unimproved” obesity states. For this analysis, we assumed values of 0.30 (compared to 0.20 in the base case) for children with unimproved obesity and 0.40 (compared to 0.60 in the base case) for those with improved obesity.

## 4 Results

Mean NHS costs after 12 months following a decision not to undertake further examination were estimated to be £1,589 while mean costs following the decision to investigate were £3,247. Mean QALYs after no further investigation were 0.24, compared to 0.47 following further investigation. The incremental cost-effectiveness ratio in the base case was therefore £7,133 per QALY. These results are shown graphically in Figures 3 and 4 below.

**Figure 3.**
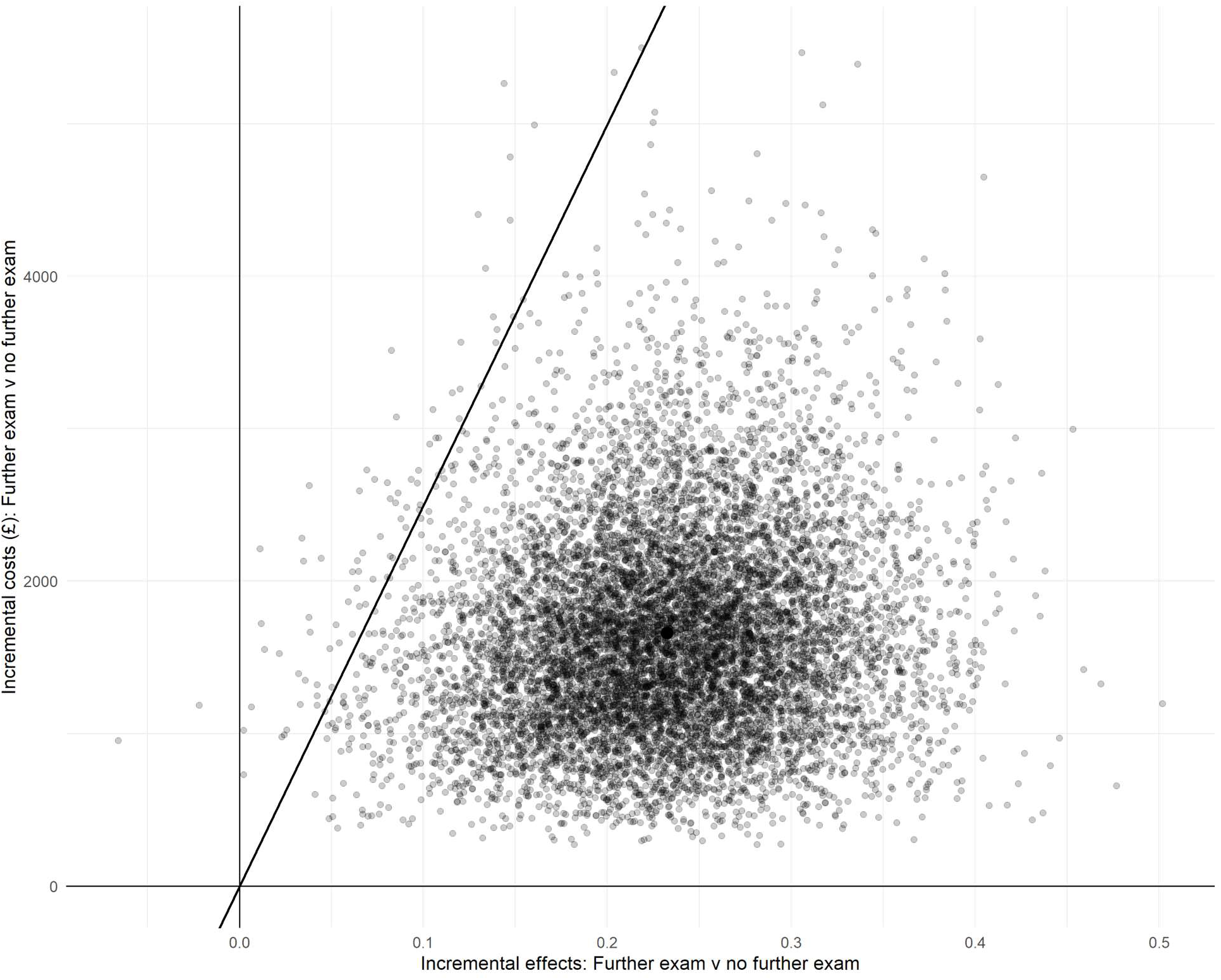
Cost-effectiveness plane (further investigation compared to no further investigation)

**Figure 4.**
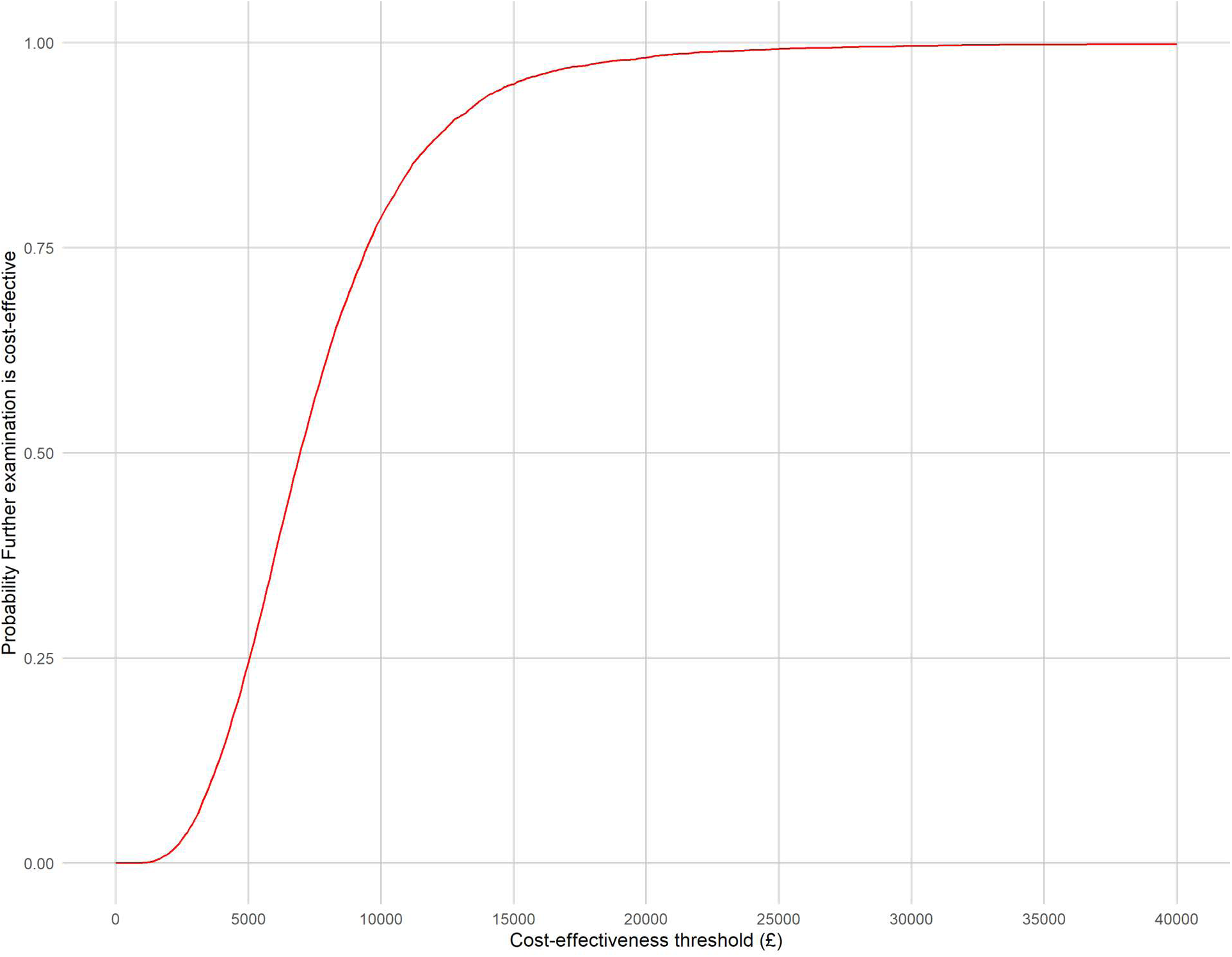
Cost-effectiveness acceptability curve (further investigation compared to no further investigation)

Figure 4 is the cost-effectiveness acceptability curve, showing the probability that further investigation is more cost-effective than no further investigation for different values of the cost-effectiveness threshold.

These results indicate that the decision to undertake further investigation is likely to be cost-effective with a probability of 99% at a cost-effectiveness threshold of £25,000 per QALY.

### 4.1 Sensitivity analysis

We conducted a deterministic sensitivity analysis to explore uncertainty in the quality-of-life (utility) parameters. In one scenario, we increased the utility assigned to the improving weight state to 0.85 (base case 0.60) while keeping the unimproved obesity utility unchanged at 0.20. In a second scenario, we tested a narrower separation between states by setting the utilities to 0.40 (improved) and 0.30 (unimproved). In the former scenario, further examination is likely to be cost-effective, being associated with an incremental cost-effectiveness ratio of £4,387. For the highly conservative scenario in which there is little difference between utility values at 12 months, the decision to undergo further examination is still cost effective, although less so, with an ICER of £28,443 and therefore below the upper threshold limit of £35,000 per QALY.

## 5 Discussion

We performed an early economic evaluation to model the cost-effectiveness of a new pathway for supporting children with BMI more than three standard deviations above mean BMI for age and sex and who may present with early onset obesity. This study therefore addressed an important and clinically underserved population: children with early-onset monogenic obesity. It is also particularly timely given the growing interest in GLP-1 receptor agonists within the NHS, highlighting the need for evidence to inform future management in this group.

The core decision modelled was whether usual care for severe obesity is maintained, or whether further examination with respect to some or all of development, growth, and genotype is undertaken. Children eligible for this pathway are not well-represented in drug trials in general and specifically in trials of GLP-1 agonists, nor is there much data on outcomes in this group. The model therefore offers a means of supporting this patient group in advance of interventional and prospective cohort studies. Our analysis of treatment decisions is likely to be relevant to different types of severe early onset monogenic obesity in children, including in patients with melanocortin-4 receptors (MC4R) variants (e.g. (36) and those with leptin deficiencies. The model suggested that further examination of children under this pathway, and possible treatment with semaglutide where indicated, is likely be cost-effective at conventional NHS cost-effectiveness thresholds.

However, there are several limitations to our analysis. The core limitation relates to the sparse availability of data on most key model parameters, encompassing probabilities, costs and QALY outcomes. When data from published literature was unavailable, particularly for parameters such as incidence, prevalence, and drug response probabilities, we relied on the clinical and laboratory expertise of the authorship group in managing the care of these children and in processing their genetic samples. Informal elicitation of parameter values from experts in this way may result in unrepresentative or inaccurate characterisations of these parameters, even after allowing for the variances attached to these parameters in probabilistic sensitivity analysis. We also did not account for any correlations between the mean values of model parameters.

There was little to no data on utility values for this group of severely obese children. For example, the utility values reported in the widely cited meta-analysis of Brown et al (33) did not necessarily correspond to the age of individuals for whom further examination might be considered in this pathway, the severity of their obesity, to a particular health-related quality of life instrument, to means of obtaining utility scores other than by direct response, or necessarily to the NHS England context. We again relied on arbitrary but potentially conservative assumptions concerning quality-of-life scores in this patient population. Conservative assumptions concerning quality of life in these children had large impacts on cost-effectiveness in deterministic sensitivity analysis.

We limited the duration of follow-up in this hypothetical model to the 12 months after the initial outpatient consultant appointment. This assumption was informed both by a lack of long-term data on the use of these drugs in children but primarily by an intention to examine the proximal, near-term outcomes of introducing this pathway. However, the use of a short duration of follow-up in the model is also likely to be somewhat conservative, given that data from adult populations indicate that longer-term use (>12 months) of semaglutide and other similar drugs is associated with sustained weight reductions and improvements in weight-related comorbidities provided that drug use is maintained (37, 38).

We did not separately model discontinuation specifically due to drug intolerance. For instance, a study by van Boxel et al (32) revealed that around 10% of children in their small prospective cohort aged 10-18 stopped taking semaglutide because of its side effects. We also did not model non-compliance in managements for patients not undergoing further examination and receiving standard of care. We also did not model any regional variation in access to specialist genomic testing services for this population.

A final issue that merits consideration is that of structural uncertainty. This refers to which features of the decision problem are represented in the model to capture relevant aspects of the both the condition and the particular clinical context and decision under investigation. Not all possible branches were modelled or parameterised. For example, we treated all abnormal R149 panel results as a single terminal node leading to setmelanotide, without distinguishing between the specific variants that may be identified. These would include principally pathogenic variants in POMC, PCSK1, and LEPR. However, the R149 panel has a low diagnostic yield in UK paediatric obesity cohorts (1/80 in Lim et al (25), with most positives being MC4R heterozygotes). Splitting the R149-abnormal branch by specific variant would add complexity without materially changing costs or effects. In any event, there is little data available to parameterize rare sub-branches that would be defined by these variants. Likewise, most children undergoing further treatment in this pathway receive semaglutide, with different genetic tests in some but not all cases establishing diagnosis rather than specific treatments. For example, the modelled panel result in phenotype 1 (R48/R263/R293/R49/R452) informs diagnostic interpretation but not treatment selection, unlike phenotype 3 where abnormal R149 results is modelled as requiring the use of setmelanotide.

## 6 Conclusion

Early economic modelling of cost-effectiveness model suggests that undertaking further investigations in this paediatric population is likely to be cost-effective for the NHS. However, there was an important lack of published evidence for many of the model parameters used to inform this conclusion, and further studies will be needed to conclusively address these issues.

## Availability of data and materials

All data and code are available at github.com/pdixon-econ/mono-obesity-decision-tree.

